# A conceptualisation and psychometric evaluation of positive psychological outcome measures used in adolescents and young adults living with HIV: a mixed scoping and systematic review

**DOI:** 10.1101/2023.07.17.23292789

**Authors:** Jermaine M. Dambi, Frances M. Cowan, Faith Martin, Sharon Sibanda, Victoria Simms, Nicola Willis, Sarah Bernays, Webster Mavhu

**Affiliations:** University of Zimbabwe: Rehabilitation Sciences Unit, Faculty of Medicine and Health Sciences, Avondale, Harare, Zimbabwe; Centre for Sexual Health and HIV/AIDS Research (CeSHHAR), Harare, Zimbabwe; Liverpool School of Tropical Medicine, Liverpool, United Kingdom; Cardiff University, Park Place, United Kingdom; London School of Hygiene & Tropical Medicine, London, United Kingdom; ZVANDIRI, Avondale, Harare, Zimbabwe; University of Sydney, Camperdown, Australia

**Keywords:** Adolescents, Global Mental Health, HIV/AIDS, psychometrics, positive psychology, systematic review: sub-Saharan Africa

## Abstract

**Introduction:** Sub-Saharan Africa bears the greatest burden of HIV/mental disorders combined. It is important to evaluate the mental health of adolescents and young adults living with HIV (AYALHIV) comprehensively by measuring both negative and positive psychological constructs. There has been a proliferation of interest in positive psychological outcome measures, but the evidence of their psychometric robustness is fragmented. This review sought to:

1. Identify positive psychological outcomes and corresponding outcome measures used in AYALHIV in sub-Saharan Africa
2. Critically appraise the psychometrics of the identified outcome measures

**Methods and analysis:** Independent and blinded reviewers searched articles in PubMed, Scopus, Africa-Wide Information, CINAHL, Psych INFO, and Google Scholar. Separate independent reviewers screened the retrieved articles. We applied a narrative synthesis to map the key constructs. The risk of bias across studies was evaluated using the COnsensus-based Standards for the selection of health Measurement INstruments (COSMIN) checklist. The quality of the psychometric properties was rated using the COSMIN checklist and qualitatively synthesised using the modified Grading of Recommendations Assessment, Development and Evaluation checklist.

**Results:** We identified 15 positive psychological constructs: body appreciation, confidence, coping, flourishing, meaningfulness, personal control, positive outlook, resilience, self-management, self-compassion, self-concept, self-efficacy, self-esteem, self-worth and transcendence that had been used to assess ALHIV. Resilience, self-concept, self-esteem, coping and self-efficacy were the most measured constructs. Construct validity and internal consistency were the properties most frequently assessed, while content validity and structural validity were assessed less often.

**Conclusions:** Few studies performed complete validations; thus, evidence for psychometric robustness was fragmented. However, this review shows the initial evidence of the feasibility of using positive psychological outcomes in low-resource settings. Instead of creating new outcomes, researchers are recommended to leverage the existing measures, adapt them for use, and, if appropriate, strive to maintain the factorial structure to facilitate comparisons.

**Registration:** PROSPERO-CRD42022325172

## Introduction

The burden of HIV in young people in low-resource settings, particularly in the sub-Saharan Africa (SSA) region, remains high [1,2]. Adolescence and early adulthood are challenging developmental stages, with the burden of navigating life challenges often even greater for adolescents and young adults living with HIV (AYALHIV) [2–4]. For instance, AYALHIV face multiple psychosocial and structural challenges, including stigma, negotiating reproductive health, socioeconomic deprivation, violence, grief, orphaning and other difficulties [1,2,4,5]. HIV has evolved into a long-term condition with a concomitant surge in comorbid non-communicable diseases [3,6]. For example, common mental disorders, including anxiety and depression, are highly prevalent in AYALHIV, with a pooled prevalence of 26.1% (95% CI 18.9-34.8) [2]. However, there are few integrated programmes combining HIV and mental health care [2–4]. Importantly, many mental health conditions that present in adulthood emerge in late adolescence and young adulthood, and effective management earlier in the life course can prevent long-term mental health difficulties [2,3]. Systematic reviews have demonstrated that access to mental healthcare by AYALHIV is associated with positive outcomes across the treatment continuum, including: increased treatment initiation, increased adherence to care, viral suppression, and reduced morbidity and mortality [3,4,7,8].

Mental health endpoints within HIV care have been traditionally conceptualised as improvements in negative psychiatric symptomatology [5,9]. For example, success in psychotherapies is invariably benchmarked against declines in rates of depression, anxiety, post-traumatic stress disorders, and other negative psychological indices [5]. However, focusing on negative indices misses the opportunity to capture mental health’s multidimensionality [10]. A holistic mental health evaluation requires a comprehensive focus on both negative and positive mental health constructs [5, 7], and recognition of this has resulted in a shift towards positive psychology, a framework that emphasises increasing human well-being and positive functioning [5,10–12]. Positive mental health interventions (PMHIs) are anchored upon the need to optimise human strengths and capabilities to improve positive outcomes such as self-esteem, resilience, hope, self-worth, social resources, and flourishing [5,9,13]. For instance, studies have shown that people living with a chronic condition (e.g., HIV) develop resilience with time [5,13]. The resilience developed in navigating the challenges of living with a chronic condition is potentially transferable into everyday functioning [13]. Positive psychology interventions (e.g., resilience-building approaches) are central to prevention health promotion and act as an entry point to stepped care for mental health problems in routine HIV care [5].

With the proliferation of PMHIs comes the need to routinely evaluate the clinical endpoints from both the clients’ and therapists’ perspectives [14]. The patient’s evaluation of their health, treatment expectations and outcomes are contingent upon the availability of validated and reliable outcome measures [9]. The last few decades have seen a proliferation of positive psychology outcome measures [15]. However, there is limited understanding of the salient positive psychological constructs linked to AYALHIV’s improved well-being and health-related quality of life. Rigorous evaluation of PMHIs is essential but limited by a lack of robust measures.

In their scoping review, Wayant et al. (2021) mapped 15 positive psychological constructs associated with increased quality of life and survival in adolescents and young adults living with cancer [16]. Well-being, personal growth, hope, meaning in life, self-esteem, vitality and optimism were the most cited positive constructs [16]; these constructs are potentially relevant to AYALHIV. Conversely, etiological differences between cancer and HIV could also lead to differences in lived experiences, resulting in differential perceptions of positive psychological constructs [16]. For instance, HIV-related stigma (often associated with issues related to HIV’s potential infectiousness) may have a more significant impact on mental health functioning in AYALHIV [12,22] when compared to the effects of cancer-related stigma [13]. It is thus critical to contextualise the impacts of positive psychological outcomes in AYALHIV.

Elsewhere, Govindasamy et al. (2021) performed a mixed-methods systematic review to explore correlates of well-being among AYALHIV in SSA to inform econometric evaluations [17]. The review showed that social support, belonging, purpose in life, and self-acceptance optimise well-being in AYALHIV [17]. Also, Orth, Moosajee and Van Wyk (2023) performed a systematic review to identify and conceptualise mental wellness in adolescents [10]. The review identified 13 concepts: life satisfaction, mental well-being, resilience, self-efficacy, self-esteem, connectedness, coping, self-control, mindfulness/spirituality, hope, sense of coherence, happiness, and life purpose. However, no psychometric evaluation of the analysed instruments was done [10,17]. Earlier work by Govindasamy et al. (2021) and Orth, Moosajee and Van Wyk (2023) provides essential insights into the broad nature of well-being conceptualisation in AYALHIV. However, the sole focus on mental well-being limits our comprehensive understanding of the spectrum of positive psychological constructs in AYALHIV living in SSA. There is need to build on earlier work and understand positive psychological constructs in HIV care for AYALHIV: such work is potentially applicable to other chronic conditions, given the multi-level impacts of HIV. Also, there is a paucity of collective evidence of the psychometric robustness of the positive psychological outcome measures used in AYALHIV. Some of the available generic outcomes may not comprehensively reflect the nuances of living with HIV [17]. Further, different investigators use varying wording to refer to the same construct; a mapping of the constructs is vital. This mixed review, therefore, sought to:

1. Identify positive psychological outcomes and corresponding outcome measures used in AYALHIV in SSA.
2. Critically appraise the psychometric properties of the identified positive psychology outcomes used in AYALHIV.

## Methods

### Overview

This mixed review was done in two sequential and complementary phases. First, a scoping review identified positive psychological outcomes used in AYALHIV in SSA and mapped the constructs onto the corresponding measures. The scoping review was performed per Preferred Reporting Items for Systematic reviews and Meta-Analyses extension for Scoping Reviews (PRISMA-ScR) guidelines - See **S1 Table** [18]. The second phase systematically evaluated the psychometric properties of the outcomes identified from the scoping review. Evaluation of outcome measures’ psychometrics was performed and reported according to the Preferred Reporting Items of Systematic Reviews and Meta-Analyses (PRISMA) guidelines [19] - See **S2 Table.** Where appropriate, we outlined specific methodological considerations unique to each phase.

### Protocol/ registration

The protocol was registered on the PROSPERO database - CRD42022325172 and was previously published [12].

### Eligibility criteria

The following criteria were applied in selecting articles:

#### Construct(s)

We included studies evaluating positive psychological constructs. We broadly defined positive psychological outcomes as any construct focusing on “… *aspects of the human condition that promote fulfilment, happiness, and flourishing…*” [20]. Positive psychology is a rapidly developing field; consequently, there is variability in the definition and conceptualization of psychological constructs [5,10–12,16,20]. We build upon operational definitions outlined by Wayant et al. in their mapping of positive psychological constructs in paediatric and adolescent/young adult patients with cancer [16]. Wayant et al.’s scoping review yielded these 15 constructs: contentment, gratitude, happiness, hope, life satisfaction, meaning in life, optimism, perseverance, personal growth, resilience, self-esteem, self-acceptance, tranquillity, vitality and well-being.

#### Study designs/interventions

For the scoping review, we included all quantitative designs, mixed methods, qualitative studies exploring the positive psychological phenomenon in AYALHIV in SSA, and grey literature (e.g., blogs and websites). For the systematic review, only quantitative designs were included. Systematic reviews, editorials, and study protocols were excluded from the scoping and systematic reviews.

#### Participants/settings

For both phases of the mixed review, we analysed all studies reporting on using and evaluating positive psychological constructs in AYALHIV (10-24 years-old) in SSA across all settings. We focused on AYALHIV as it is the group with the greatest burden of HIV globally [17]. We anticipated that some studies contained data on AYALHIV and other age bands (e.g., children and middle-aged adults). In such cases, an article was considered for review if the average age was within the 10-24 years range or if over 50% of the participants were AYALHIV.

#### Language

We restricted the analysis to articles published in English for both phases of the mixed review. We did not have the resources to analyse articles published in other languages.

### Information sources

Peer-reviewed articles were searched/retrieved from these electronic databases: PubMed, Scopus, Web of Science, Africa-Wide Information, CINAHL, PsychInfo, and Google Scholar. Databases were searched from inception through February 28^th^, 2023. Where only an abstract was available online, and information regarding psychometrics was neither clear nor available from the text, an attempt to contact the lead author was made, requesting the full article to ensure literature saturation and a truthful rating. The article was excluded from the review if there was no response in two weeks following three email reminders. We also reviewed grey literature using the Google Scholar search engine to search potential databases such as university databases, research reports, pre-prints, newsletters and bulletins, policy briefs, guidelines, and conference proceedings for articles. For completeness, we also performed both backwards and forward searches of the reference lists of identified articles and databases, respectively. Finally, we also contacted experts implementing PMHIs to check for articles we may have missed during the literature searches.

### Search strategy

For the scoping review, as an illustration, articles in CINAHL were searched using the AND Boolean logic operators, i.e., 1 AND 6 AND 9 AND 12 (**S3 Table**). The search strategy was amended for the systematic review component to include additional constructs identified through the scoping review.

### Data management

Retrieved articles were imported into a password-protected Mendeley reference manager. The articles were also synchronised onto Mendeley and Dropbox cloud storage platforms and backed-up onto a password-encrypted external hard drive. All collaborators had full access/administrative privileges to the shared Dropbox folder for the present systematic review. A trail/history of the electronic searches was also saved on users’ PubMed, Scopus, and EBSCOhost accounts. We also printed summaries of all the searches to enhance the data capturing of the search records.

### Data collection process

The data collection process was conducted in three stages, i.e., article retrieval, screening, and data extraction. These processes were invariably similar for the scoping and systematic review phases. Here, we describe these processes and highlight, where appropriate, differences in the two phases of the review.

#### Article retrieving

Two researchers (SS & JMD) independently searched articles using a pre-defined search strategy. The lead author (JMD) then imported the searches into Mendeley and removed duplicates.

#### Screening

Upon completion of article retrieval, another set of independent researchers (SB & WM) screened the articles by title and abstract using Rayyan software [21]. To increase methodological rigour, both researchers independently reviewed all retrieved articles, including documenting reasons for exclusion. Rayyan software automatically collates the number of hits assigned different ratings by reviewers. Discrepancies were resolved through discussion, and where consensus was not reached, a more senior researcher (WM) made the final decision. JMD and SS then perform backwards and forward citation searches to identify other potential articles. Two senior researchers (FMC & WM) reviewed the list of identified articles afterwards to check for the completeness of the searches.

#### Data extraction

Once searches were finalised, two researchers (FM and NW) retrieved the full articles and independently extracted data from articles meeting the inclusion criteria. Data extraction was performed in duplicate. Disagreements during data extraction were resolved through consensus, and more senior researchers (FMC & WM) made the final decisions if any impasses occurred. For both phases of the review, we extracted the following information per study: research setting and design, study sample and participants’ demographics. For the systematic review component, we extracted information on the mode of administration, the number of items, descriptions of domains, scoring and interpretation of scores and whether measures were free to use or required a license fee or other payment.

### Charting/Outcomes and prioritisation

Qualitative conceptualisation of positive psychological constructs and the appraisal of psychometric properties of the identified outcome measures were the primary outcomes of the scoping and systematic review phases, respectively. For the systematic review, the clinical utility of the identified outcome measures was the secondary outcome. See **S4 Table** for operational definitions of psychometric properties for the systematic review component [22,23].

### Risk of bias-individual studies

The scoping review aimed to understand the conceptualisation of AYALHIV’s positive psychological constructs. Consequently, we did not perform any risk of bias (RoB) assessments. However, the systematic review component aimed to synthesise the evidence of psychometric robustness, necessitating RoB assessment. We used the revised COnsensus-based Standards for the selection of health Measurement INstruments (COSMIN) checklist to assess the RoB across studies retrieved for psychometric evaluation [22,23]. The COSMIN methodology consists of three steps. The checklist consists of methodological benchmarks for ten (10) psychometric properties, which are categorised into three major groups, i.e. content validity (e.g. patient-reported outcome measure development), internal structure (e.g. structural validity), and other psychometrical properties (e.g. criterion validity) [22,23]. Each psychometric property is rated using a pre-set criterion, and using the principle of “worse score counts”, the lowest rating is ascribed as the overall methodological quality rating [23]. Methodological quality is rated on a four-point Likert scale, i.e. “inadequate”, “doubtful”, “adequate”, and “very good“; the higher the rating, the lower the risk of bias [22,23]. We anticipated that not all details might be recorded for the retrieved articles, especially for studies whose primary aim was not psychometric evaluation. We, therefore, contacted the corresponding author to achieve the most truthful rating of the psychometric property to decrease bias during analysis.

### Quality of psychometric properties and data extraction

The quality of psychometrical properties was evaluated using an updated, hybrid checklist based on previous work by Terwee et al. [24] and Prinsen et al. [25] **(See S5 Table).** Each psychometric property was rated as; sufficient (+), insufficient (–), or indeterminate (?) [22]. Positive ratings represent high-quality psychometrics [22].

### Best evidence synthesis

We applied a narrative synthesis to map the key “themes/constructs” emerging from the scoping review. We mapped the constructs to corresponding outcome measures; this mapping exercise subsequently guided the psychometric evaluation. The collective evidence per psychometric property per outcome was synthesised using the modified Grading of Recommendations Assessment, Development, and Evaluation (GRADE) checklist [26]. The modified GRADE checklist was then used to collate the RoB results and the quality of psychometric ratings to qualitatively synthesise/summarise the quality of evidence per psychometric property across studies. A meta-analysis was not possible given the heterogeneity of outcome measures retrieved. The quality of evidence per psychometrical property was classified as very low, low, moderate or high [26] - **See S6 Table**.

### Patient and Public Involvement statement

We worked collaboratively with AYALHIV during data collection and dissemination. AYALHIV representatives previously trained and involved in systematic reviews assisted with article screening. This review is attached to ongoing work in which AYALHIV are collaboratively engaged. It is part of a larger study to explore various constructs to understand how they improve AYALHIV’s health outcomes. We have recruited AYALHIV to serve as a Youth Expert Panel (YEP). The YEP functions as both a guide to the study/research process and an additional group of analysts and discussants to examine the emerging analysis and findings. We also co-created the dissemination plans; for instance, adolescents and young adults with lived experiences were involved in co-developing output animation and contributing to the project blogs, amongst other dissemination activities.

### Ethics and dissemination

No ethical approvals were needed as this is a literature review. The mixed review maps and appraise the collective evidence of the psychometric robustness of positive psychological outcomes used in AYALHIV. The review builds on recommendations of systematic reviews on objectively measuring positive psychological constructs across diverse populations. This is important given the need to use valid and reliable outcomes in understanding the positive effects of living with HIV. The review also assisted in identifying psychometrically robust outcomes to inform an item bank to adapt a context-specific outcome measure for AYALHIV in low-resource settings. For example, we consolidated all self-esteem outcome measures and categorised items from multiple outcomes into common factors/“themes”. The outputs collectively informed the development, implementation, and evaluation of a bespoke positive mental health intervention for AYALHIV, hence a need for a multimodal dissemination plan to reach multiple stakeholders. In addition to publishing the outcomes in a peer-reviewed journal, we disseminated the outcomes through social media, policy briefs and blogs.

## Results

The results are presented in two parts. First, we present the mapping of constructs identified from the analysed studies from the scoping review component. The second part presents the qualitative synthesis of standardized outcome measures analysed in the systematic review.

### Study selection

We retrieved 6437 studies, of which 1 679 were duplicates. After de-duplication, 4748 articles were screened by title and abstract; of these, 4050 were assessed for eligibility. Six (60) articles met the full criteria and were analysed in this review. See **S7 Figure** for the PRISMA flowchart.

### Description of study participants and settings

Slightly over half of the outcomes were developed in high-income settings (15/29), with 19 studies conducted in urban areas, six (6) in rural settings and four (4) in both urban and rural localities. 13/29 (45%) of the studies were cross-sectional, with 20/29 (69%) of the studies published between 2018 and 2023 **- See Error! Reference source not found..**

**Table 1:** Description of study participants and settings.

| Construct | Name of tool | Country; setting | Design | Participants, Sample size | Age (years) |
| --- | --- | --- | --- | --- | --- |
| Body Appreciation | Body appreciation scale 2 (BAS-2) | South Africa; Urban Primary care clinics | Cross-sectional. | YPLHIV (N=76) | Range:15–24; Mean (SD): 19.4 (2.6) |
| Confidence | Ad hoc | Zimbabwe; rural primary healthcare clinics | RCT | ALHIV, (N=94) | Range: 10-15 |
| Coping | Psychological Adjustment to Illness Scale Self Report (PAIS-SR) | Nigeria; urban | Quasi-experimental | Non-Disclosed YPLHIV, ( N=19) | Range: 15–29 |
| Coping | Acceptance of illness scale (AIS) | Nigeria; Rural and Urban HIV treatment centres | Mixed methods | Pregnant women living with HIV (N=840) | Range: 22-46 |
| Coping | Ad hoc | South Africa; primary care clinics | RCT | Women living with IV (N=143) | Range: 18-50: Mean (SD) control: 28.4 (6.3) : Mean (SD) intervention: 30.6 (5.8) |
| Flourishing | Flourishing Well Being Scale (FWBS) | South Africa; households; households | Cross-sectional | AGYW living with HIV (N=568) | Range: 10–24 |
| Meaningfulness | HIV meaningfulness scale (HIVMS) | Nigeria; Rural and Urban HIV treatment centres | Mixed methods | Pregnant women living with HIV (N=840) | Range: 22-46 years |
| Personal Control | Mastery Scale (MS) | Ghana; Urban-Hospital-based clinic | Cross-sectional | PLHIV in Ghana & USA (N=55 Ghana) | Range 15-49 |
| Positive Outlook | Positive Outlook-Individual Protective Factors Index (PIPFI) | Uganda; Urban-Community clinic | Retrospective cohort study | Children living with HIV, exposed and unexposed to HIV (N=165) | Range: 6-18: Mean (SD): 10.8 (3.5) |
| Resilience | Child Youth Resilience Measure-12 (CYRM-12) | South Africa; Urban-public health ART clinics | Cross-sectional | ALHIV, (N=385) | Range 13-18: Median (IQR): 15 (14-16) |
| Resilience | Connor-Davidson Resilience scale (CDRS-25) | South Africa: Rural-Community-based | Cross-sectional survey | YPLHIV (N=334) | Range: 12-24 : Median (IQR): 21 (16 to 23) |
| Resilience | Connor-Davidson Resilience scale (CDRS-25) | South Africa; Rural-public healthcare facilities | Cross-sectional survey | YPLHIV (N=359) | Range: 12-24: Median (IQR): 21 (16- 23) |
| Resilience | Connor-Davidson Resilience scale (CDRS-10) | South Africa; Households | Cross-sectional | AGYW living with HIV (N = 568) | Range 10–24 |
| Self-Management | Adolescent HIV Self-Management Scale (AdHIVSM) | Lesotho; Urban – hospital and Youth centre | Cross-sectional survey | AYLWH (N = 183) | Range: 15–25: Median (IQR): 22 (4) |
| Self-Management | Adolescent HIV Self-Management Scale (AdHIVSM) | South Africa; Urban – healthcare facilities | cross-sectional | ALHIV (N=385) | Range: 13–18 Median (IQR): 15 (14-16) |
| Self-Management | Adolescent HIV Self-Management Scale (AdHIVSM) | South; Africa; Urban – healthcare facilities | mixed method | ALHIV (N=385) | Range: 13-18: Median (IQR): 15 (14 – 16) |
| Self-compassion | Self-compassion scale (SCS) | Nigeria; Rural and Urban HIV treatment centres | Mixed methods | Pregnant women living with HIV (N=840) | Range 22-4 |
| Self-Concept | Beck Youth Self-Concept Scale (BYSCS) | South Africa; Urban-HIV clinics | Cohort | YLP HIV (N=203), HIV-U (N=44) | Range 9–11: Median (IQR) YLP HIV: 10.7 (9.9 – 11.4) Median (IQR): HIV-U: 10.3 (9.7-11.1) |
| Self-Concept | Beck Youth Self-Concept Scale (BYSCS) | South Africa; Urban-Public sector healthcare service | prospective cohort study baseline data | PHIV+ adolescents (N=204), Control (N=44) | Range 9-11: HIV+ mean = 10.4 SD 0.9; controls Mean = 10.4 SD = 1.1 |
| Self-Concept | Beck Youth Self-Concept Scale (BYSCS) | South Africa; Urban – Research Centre | Cohort study | ALHIV (N=122) | Range 12–15 PHIV+ (Mean SD): 13.5 (1.0); Controls (Mean SD):13.8 (1.2) |
| Self-Concept | Tennessee Self-concept Scale-2 (TSCS-2) | Uganda; Rural – Public schools | longitudinal study | AIDS orphaned adolescents (N=268) | Range 11-17:Mean (SD): 13.7 (1.3) |
| Self-Concept | Tennessee Self-concept Scale-2 (TSCS-2) | South Africa; Urban-HIV clinics | Longitudinal study | Perinatally infected YLHIV (N=37) | Range: 9-14; Mean (SD): 11.6 (1.7) |
| Self-Concept | Tennessee Self-concept Scale-2 (TSCS-2) | South Africa; Urban – Paediatric HIV clinics and public hospitals | RCT baseline data | PHIV (N=177) | Range: 9-14; Mean (SD): 11.68 (1.42) |
| Self-Efficacy | Self-Efficacy Questionnaire for Children | Tanzania; Urban – weekly paediatric clinic | Pilot randomized waitlist-controlled trial | ALHIV (N=48) | Range: 14-18; Mean (SD): 15.7 (1.4) |
| Self-Efficacy | Self-Efficacy for Managing Chronic Disease 6-Item Scale (SE-6-Xhosa) | South Africa; Urban-Community Health Centre | Cross-sectional | Xhosa women with HIV (N = 229) | Range 18-40; Mean (SD): 30.7 (4.8) |
| Self-Efficacy | HIV-Adherence self-efficacy assessment survey (HIV-ASES) | Kenya; Urban- HIV care and treatment outpatient clinic | Cross-sectional | ALHIV (N=82) | Range 16-19; Median (IQR): 17 (16 – 18) |
| Self-Efficacy | Ad hoc | South Africa; Urban – primary schools | Mixed Methods | Female adolescents (N=382) | Range: 11-16 |
| Self-Efficacy | Ad hoc | South Africa; Urban-Primary care clinics | Pilot Interventional study | Women living with HIV (N=120) | Range: 18-50; Mean (SD) Control: 28.4 (6.3) ;Mean (SD) Intervention: 30.6 (5.8) |
| Self-Efficacy | Ad hoc | Eswatini;<br>Rural and<br>Urban HIV<br>care and<br>treatment<br>facilities | Cross-sectional | ALHIV (N=40) | Mean (SD):<br>15.5 (1.6) |
| Self-Efficacy | Ad hoc | Uganda; and<br>Kenya;<br>Facility-based | Cross-sectional | ALHIV (N=582) | Mean (SD):<br>14.6 (1.4) |
| Self-esteem | Rosenberg Self-esteem Measure<br>(RSEM-10) | Nigeria;<br>Urban-HIV<br>support care<br>centre | Cross-sectional | HIV positive<br>adults | Range: 18-62<br>;Mean (SD):<br>30.9 (11.4) |
| Self-esteem | Rosenberg Self-esteem Measure<br>(RSEM-10) | Ghana;<br>Urban-Hospital-based clinic | Cross-sectional | PLHIV in Ghana<br>& USA (N= 55<br>Ghana) | Range 15-49 |
| Self-esteem | Rosenberg Self-esteem Measure<br>(RSEM-10) | South Africa;<br>Urban –<br>primary<br>schools | Mixed<br>Methods | Female<br>adolescents (N =<br>382) | Range: 11-16 |
| Self-esteem | Rosenberg Self-esteem Measure<br>(RSEM-10) | South Africa:<br>Rural-Community<br>based | Cross<br>sectional<br>survey | YPLHIV (N=334 | Range: 12-24;<br>Median (IQR):<br>21 (16-23) |
| Self-esteem | Rosenberg Self-esteem Measure<br>(RSEM-10) | Namibia;<br>Rural –<br>Health<br>Centre | Exploratory<br>design | PLWHA (N=124) | Range: 13-74;<br>Mean (SD):<br>31.8 (10.9) |
| Self-esteem | Rosenberg Self-esteem Measure<br>(RSEM-10) | South Africa;<br>Rural-public<br>healthcare<br>facilities | Cross<br>sectional<br>survey | YPLHIV (N=359) | Range: 12-24<br>Median (IQR):<br>21 (16-23) |
| Self-esteem | Rosenberg Self-esteem Measure<br>(RSEM-10) | South Africa;<br>urban | Cross-sectional. | YPLHIV (N=76) | Range:15-24;<br>Mean (SD):<br>19.4 (2.6) |
| Self-esteem | Rosenberg Self-esteem Measure<br>(RSEM-10) | Tanzania;<br>Urban –<br>weekly<br>paediatric<br>clinic | pilot<br>randomized<br>waitlist-controlled<br>trial | ALHIV (N=48) | Range: 14- 18<br>Mean (SD):<br>15.7 (1.4) |
| Self-esteem | Rosenberg Self-esteem Measure<br>(RSEM-10) | *Ghana*;<br>Urban-hospital | Cross-sectional<br>study | ALHIV (N=139) | Range: 13-19;<br>Mean<br>(SD):16.6<br>(1.8) |
| Self-esteem | Rosenberg Self-esteem Measure<br>(RSEM-10) | Kenya;<br>Urban- HIV<br>care and<br>treatment<br>outpatient<br>clinic | Cross-sectional | ALHIV (N = 82) | Range 16-19;<br>Median (IQR):<br>17 (16-18) |
| Self-esteem | Rosenberg Self-esteem Measure<br>(RSEM-10) | Uganda and<br>Kenya;<br>Facility-based | Cross-sectional | ALHIV (N=582) | Mean (SD):<br>14.6 (1.4) |
| Self-esteem | Modified<br>Rosenberg Self- | South Africa;<br>Urban – | Mixed<br>Methods | Female<br>Adolescents (N =<br>382) | Range: 11-16 |
|  | esteem Measure (RSEM-8) | primary schools |  |  |  |
| Self-esteem | Tennessee Self-concept Scale-2 (TSCS-2) | Uganda; Rural – Public schools | Longitudinal study | AIDS orphaned adolescents (N=268) | Range 11-17; Mean (SD): 13.7 (1.3) |
| Self-esteem | Self-esteem-Hare Area-specific self-esteem scale | Uganda; Urban-Community clinic | Retrospective cohort study | children with and without perinatal HIV Infection/ exposure (N=165) | Range: 6-18; Mean (SD): 10.8 (3.5) |
| Self-esteem | Ad hoc | South Africa, Primary care clinics | RCT | Women living with HIV (N=143) | Range: 18-50 |
| Self-esteem | Ad hoc | Uganda and South Africa- Facility-based | Cross-sectional | Adults receiving palliative services (N=285) | mean (SD): 40.1 (12.8) |
| Self-worth | Ad hoc | Zimbabwe; Rural – Primary Healthcare Clinics | RCT | ALHIV (N=94) | Range: 10-15 |
| Transcendence | Missoula Vitas Quality of Life Index (MVQOLI) Transcendent Subscale | Uganda and South Africa; Rural to urban non-profit and state service, palliative care services and voluntary sector hospice service | Cross-sectional | Adults receiving palliative services (N=285) | mean (SD): 40.1 (12.8) |

### Qualitative mapping of positive constructs - scoping review results

**Figure 1:**
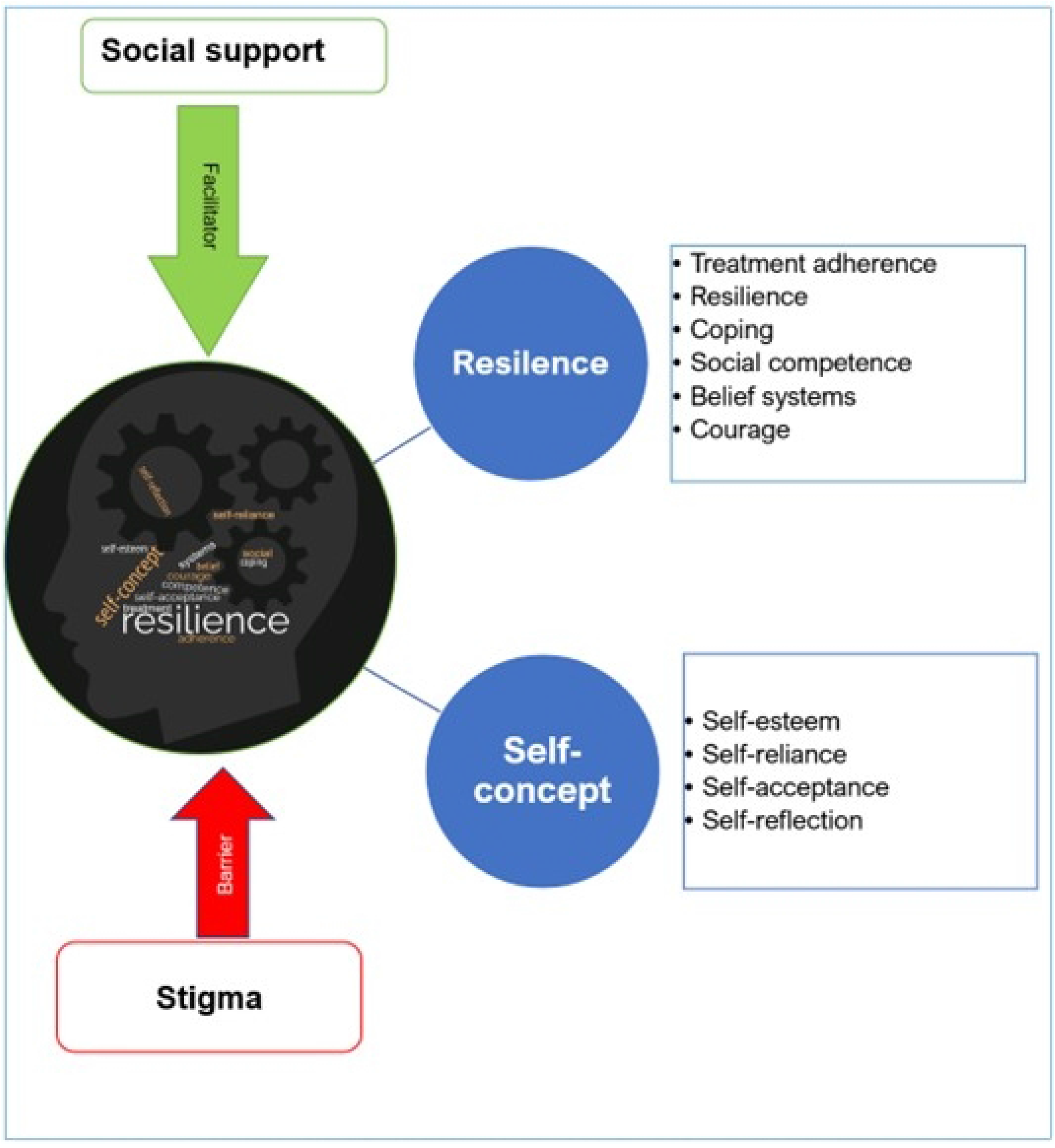
Qualitative mapping of positive psychological constructs.

From the qualitative studies, six positive psychological constructs emerged, i.e., courage, self-reliance, self-esteem, self-acceptance, resilience, and coping. Self-concept was conceptualised as an interaction of self-esteem, self-reliance, self-acceptance, and self-reflection. Self-concept is central to positive functioning; for example, high self-esteem is essential for living with HIV. The studies also suggested that resilience was crucial to coping with the demands of living with a chronic condition. Sociocultural belief systems shape resilience and are essential for treatment adherence. Social support optimised positive mental health function, with participants citing support from several sources (e.g., family and peers). Lastly, stigma and fear of disclosure were seen as the most significant barriers to positive psychological functioning - See **S8 Table** for further details.

### Quantitative mapping of positive constructs - systematic review results

#### Description of outcome measure characteristics

We retrieved 36 outcome measures spanning 15 positive psychological constructs. Resilience, self-concept, self-esteem, coping and self-efficacy were the most commonly reported constructs, as visually depicted in Figure 2. The item range for the outcomes was 5-45, with 19/36 (53%) scored on a 5-point Likert scale and most (29/36, 81%) available for free/without payment. However, a few outcome measures (11/36, 31%) had scoring instructions available - **See S9 Table**.

**Figure 2:**
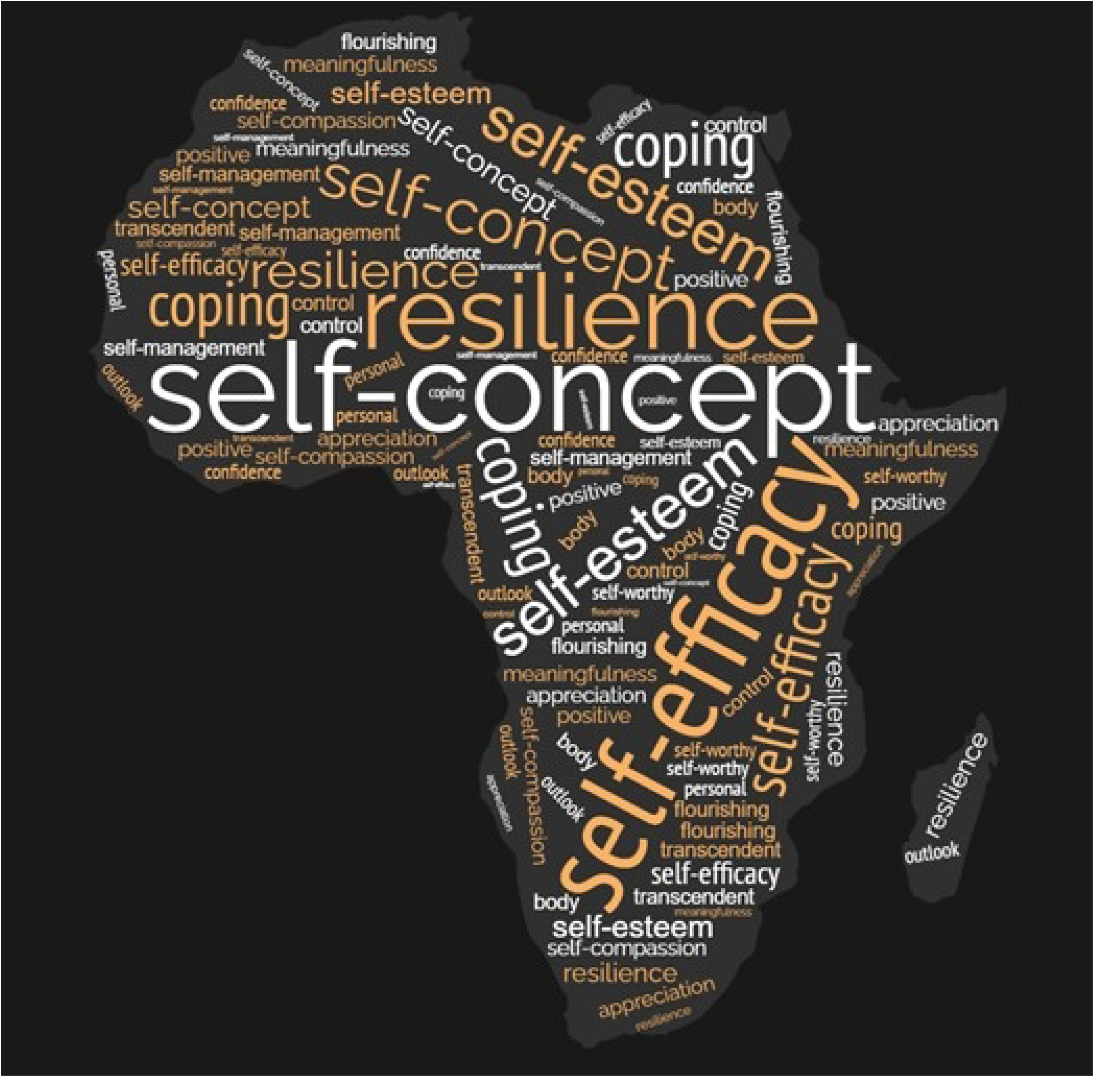
Constructs Map.

#### Results of individual outcomes sorted by construct

A description of the different outcome measures is presented subsequently; results are arranged alphabetically per construct. In **Table 2** we present methodological quality/RoB assessment ratings, with **Table 3** outlining the collation of quality of outcomes and best evidence synthesis.

**Table 2:**
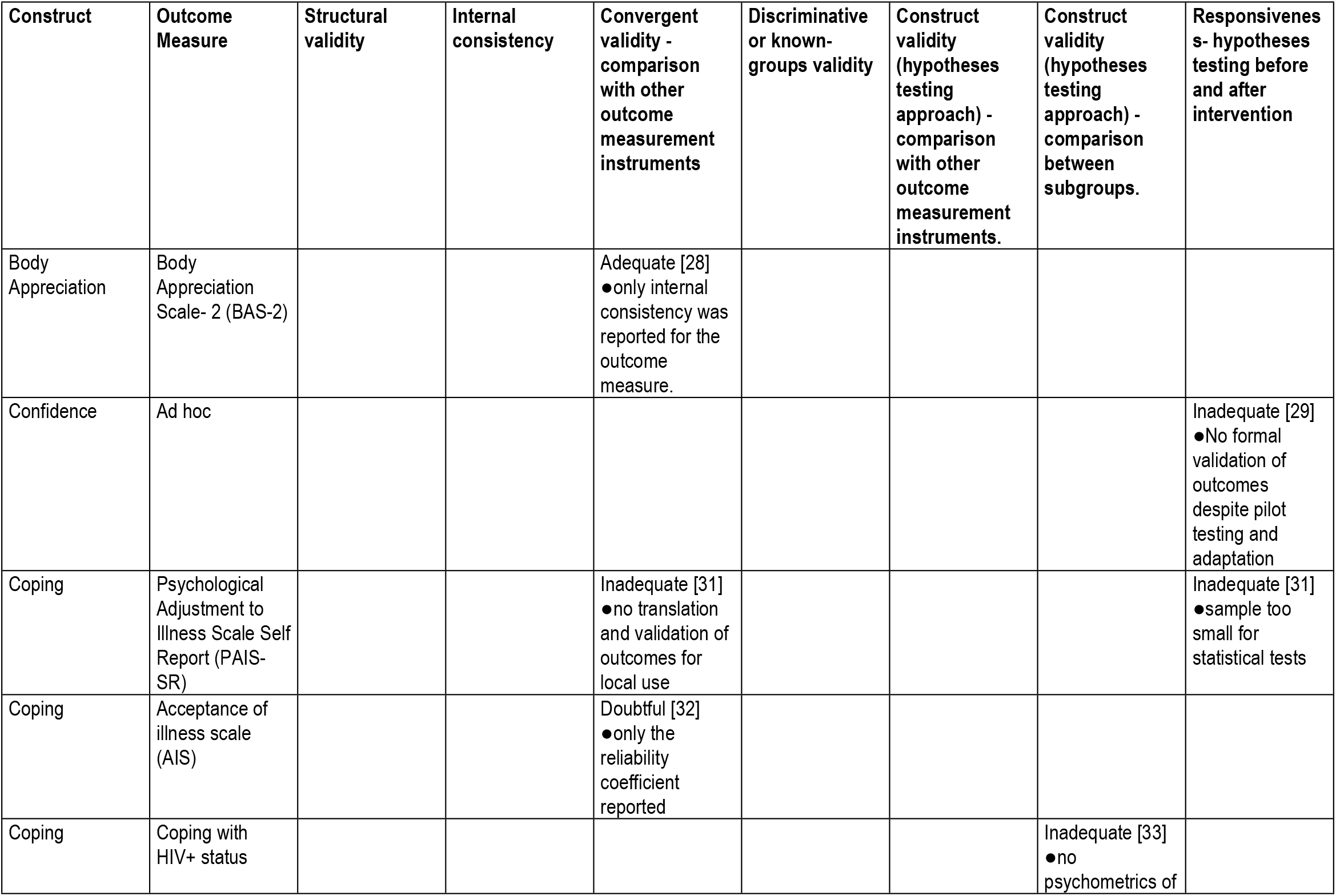
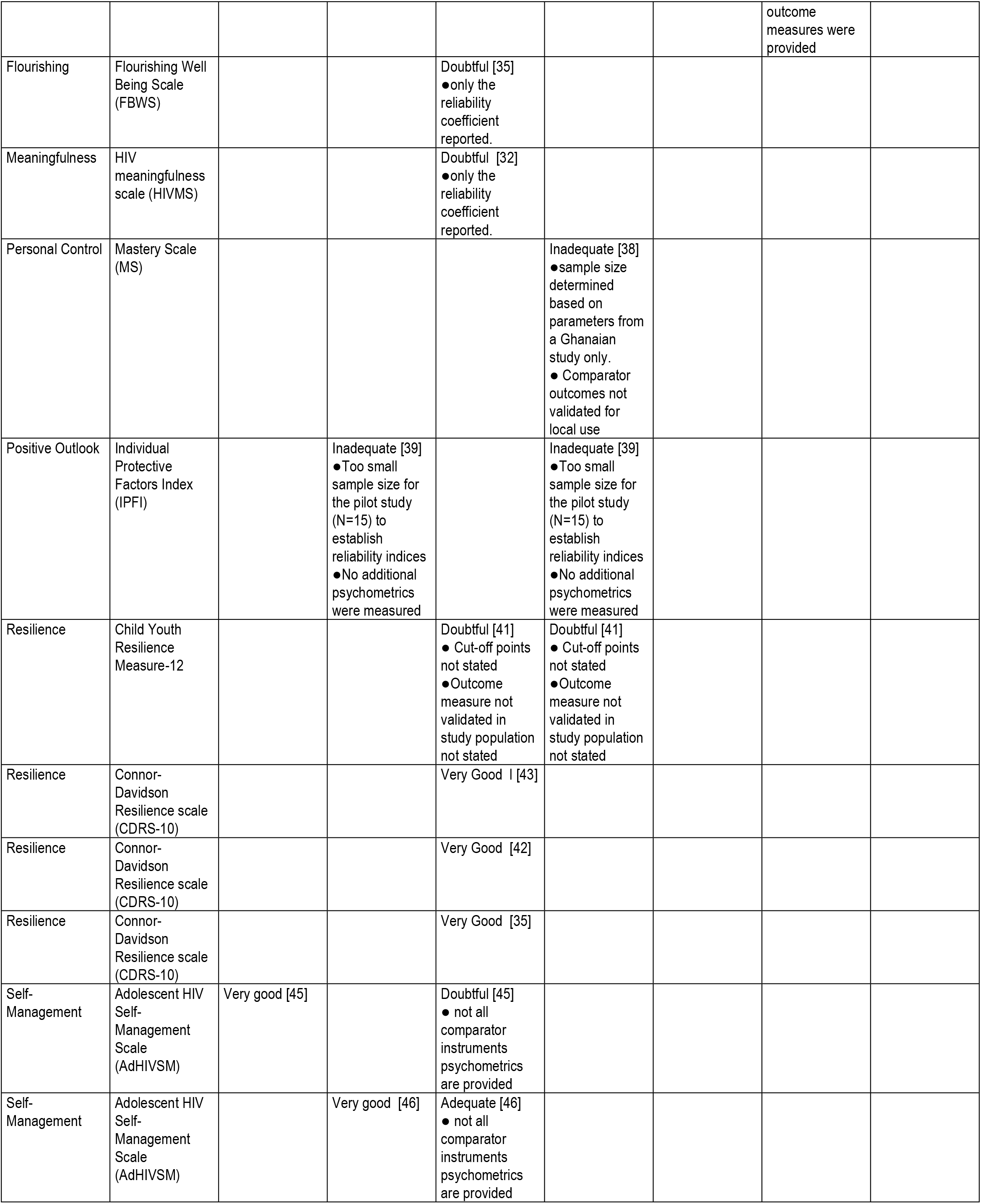
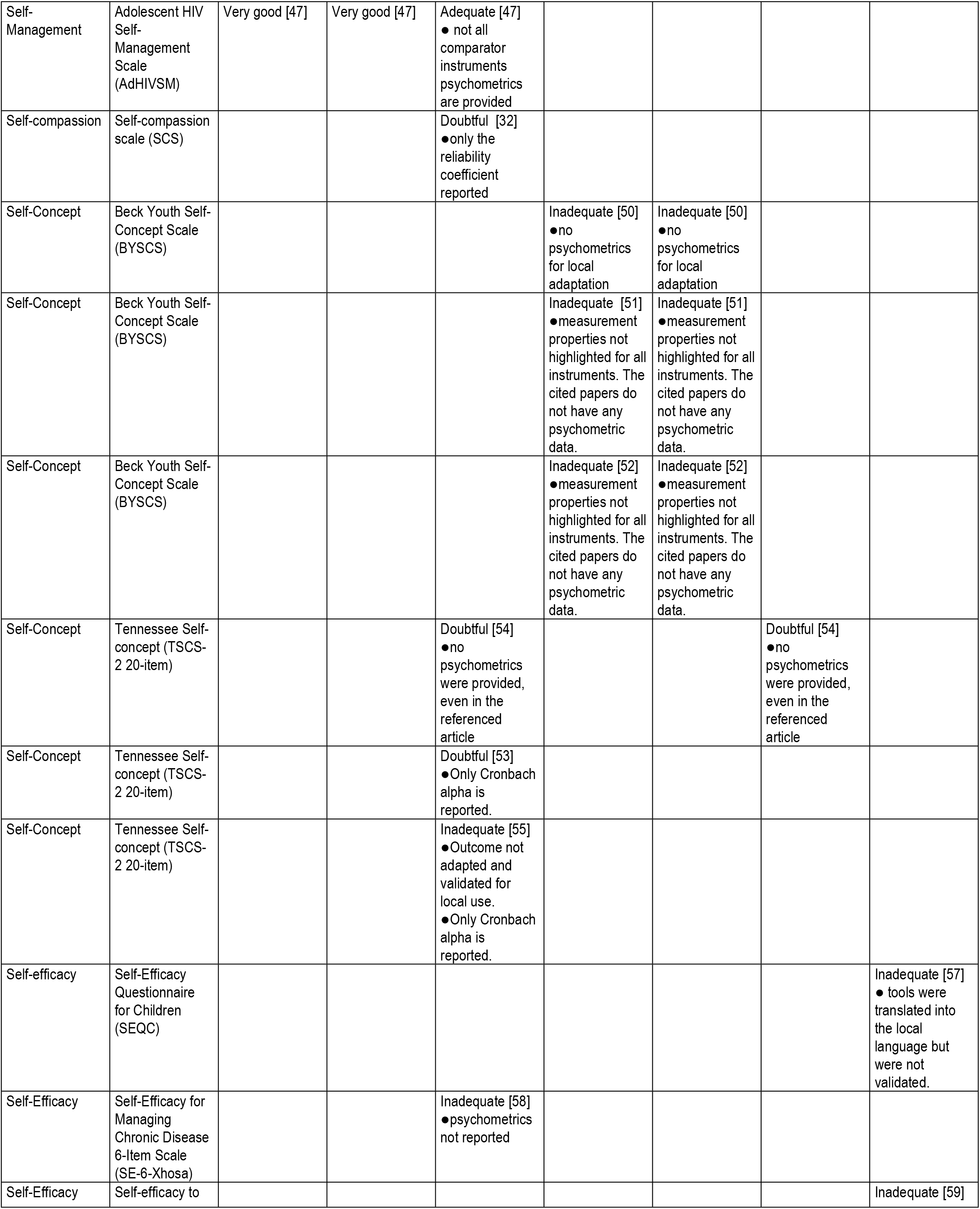
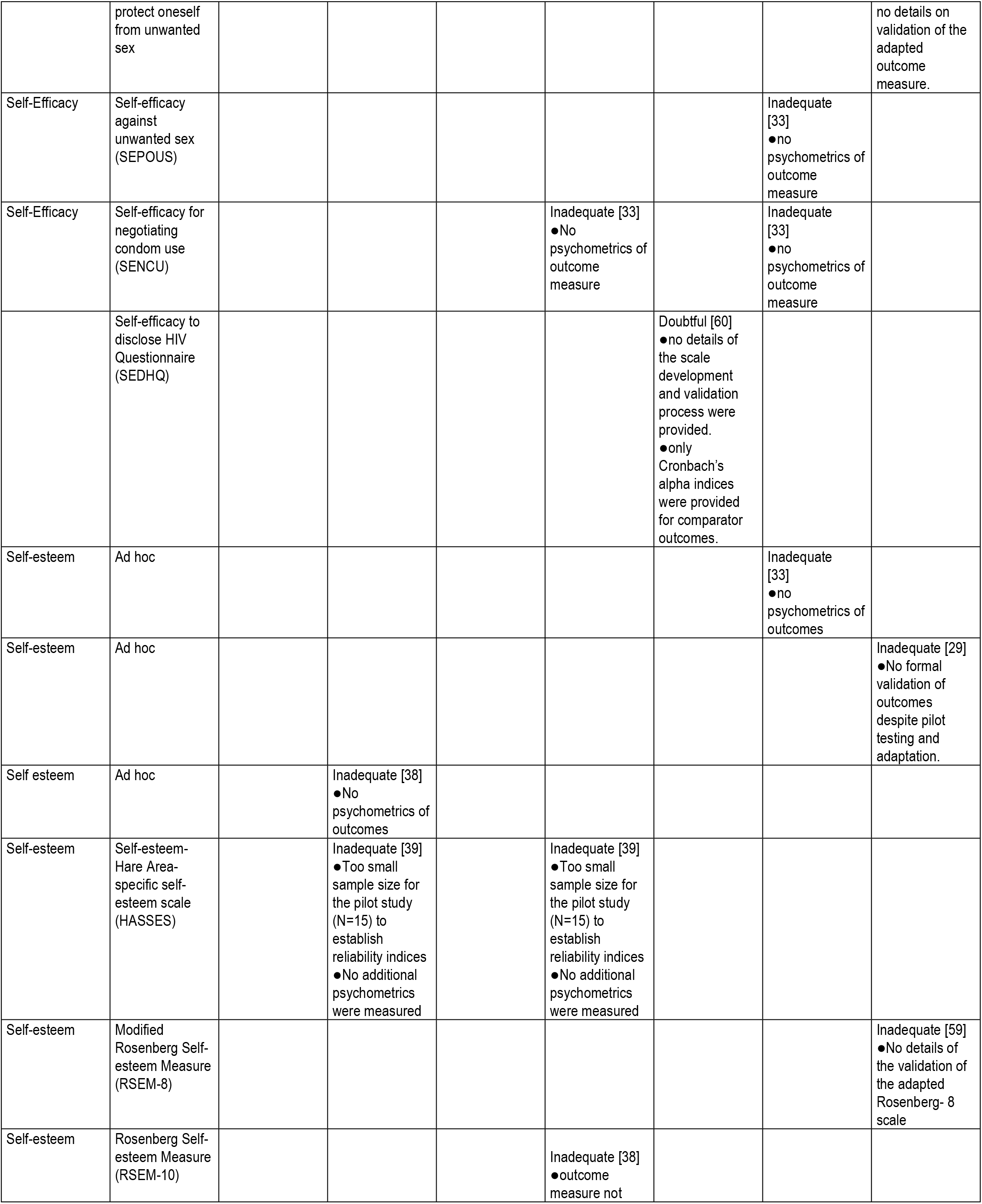
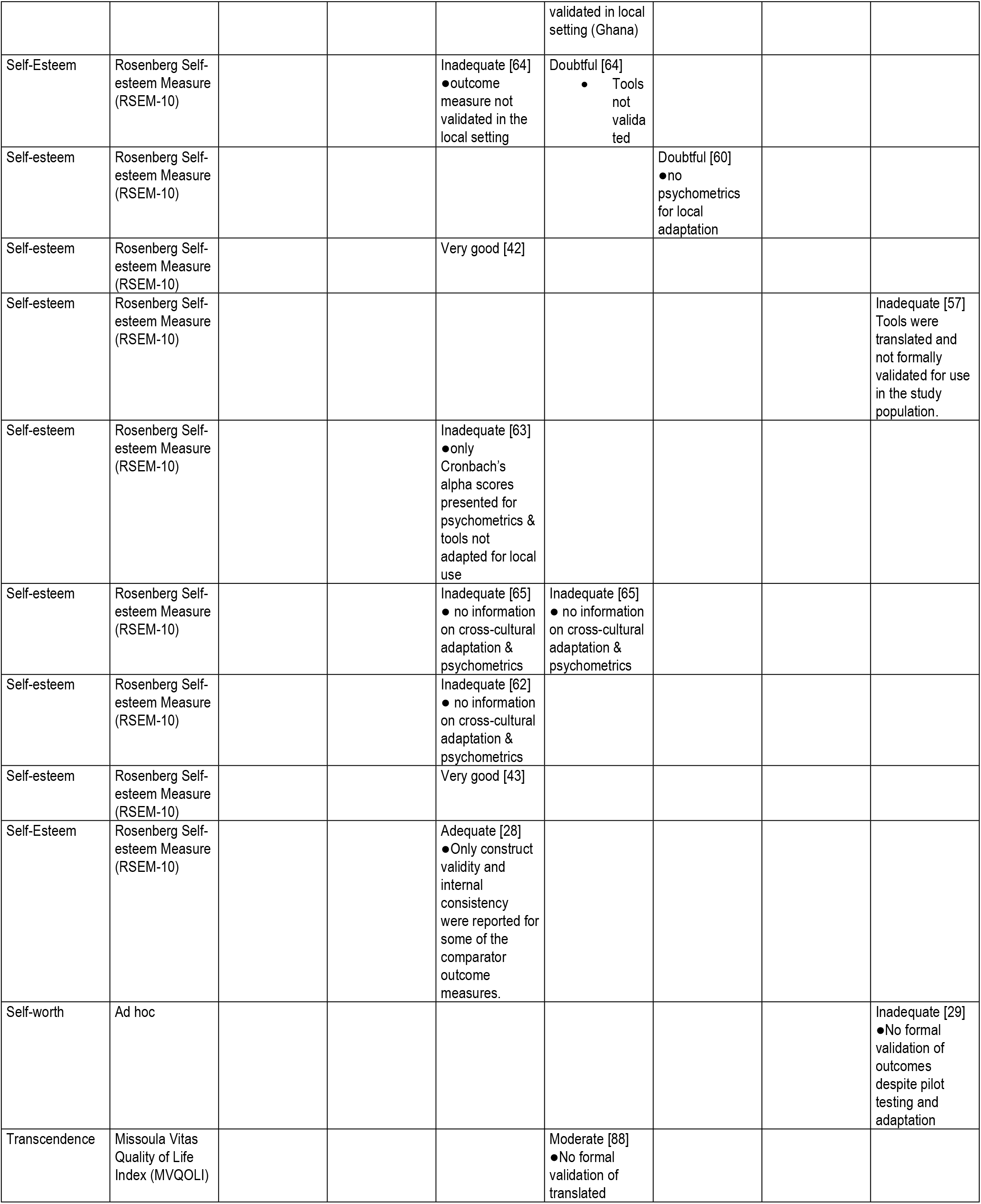
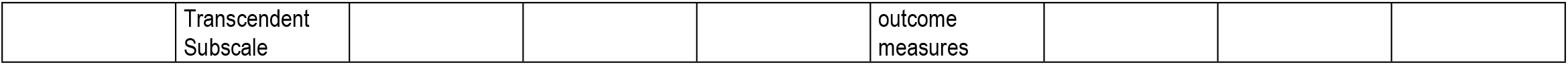
Methodological ratings.

**Table 3:** Quality of psychometrics/evidence synthesis.

| Construct | Outcome Measure | Structural validity | Internal consistency | Convergent validity - comparison with other outcome measurement instruments | Discriminative or known-groups validity | Construct validity (hypotheses testing approach) - comparison with other outcome measurement instruments. | Construct validity (hypotheses testing approach) - comparison between subgroups. | Responsiveness - hypotheses testing before and after intervention |
| --- | --- | --- | --- | --- | --- | --- | --- | --- |
| Body Appreciation | Body appreciation scale 2 (BAS-2) |  |  | Methodology quality: Adequate<br>Psychometric quality: +<br>**Quality of evidence:** Moderate [28] |  |  |  |  |
| Confidence | Ad hoc |  |  |  |  |  |  | Methodology quality: Inadequate<br>Psychometric quality: +<br>**Quality of evidence:** Very low [29] |
| Coping | Psychological Adjustment to Illness Scale Self Report (PAIS-SR) |  |  | Methodology quality: Inadequate<br>Psychometric quality: +<br>**Quality of evidence:** Very low [31] |  |  |  | Methodology quality: Inadequate<br>Psychometric quality: +<br>**Quality of evidence:** Very low [31] |
|  | Acceptance of illness scale (AIS) |  |  | Methodology quality: Doubtful<br>Psychometric quality: +<br>**Quality of evidence:** Low [32] |  |  |  |  |
|  | Coping with HIV+ status |  |  |  |  |  | Methodology quality: Inadequate |  |
|  |  |  |  |  |  |  | Psychometric quality: -<br>**Quality of evidence:**<br>Very low [33] |  |
| Flourishing | Flourishing Well Being Scale (FBWS) |  |  | Methodology quality: Doubtful<br>Psychometric quality: -<br>**Quality of evidence:** Low [35] |  |  |  |  |
| Meaningfulness | HIV meaningfulness scale (HIVMS) |  |  | Methodology quality: Doubtful<br>Psychometric quality: +<br>**Quality of evidence:** Low [32] |  |  |  |  |
| Personal Control | Mastery Scale (MS) |  |  |  | Methodology quality: Inadequate<br>Psychometric quality: ?<br>**Quality of evidence:** Very low [38] |  |  |  |
| Positive Outlook | Individual Protective Factors Index (IPFI) |  | Methodology quality: Inadequate<br>Psychometric quality: +<br>**Quality of evidence:** Very low [39] |  | Methodology quality: Inadequate<br>Psychometric quality: -<br>**Quality of evidence:** Very low [39] |  |  |  |
| Resilience | Child Youth Resilience Measure-12 |  |  | Methodology quality: Doubtful<br>Psychometric quality: +<br>**Quality of evidence:** Low [41] | Methodology quality: Doubtful<br>Psychometric quality: +<br>**Quality of evidence:** Low [41] |  |  |  |
|  | CDRS-25 |  |  | **Study 1** [43]:<br>Methodology quality: doubtful<br>Psychometric quality: +<br>**Study 2** [42]:<br>Methodology quality: very good<br>Psychometric quality: -<br>**Overall Quality of evidence:** Moderate |  |  |  |  |
|  | CDRS-10 |  |  | Methodology quality: Doubtful |  |  |  |  |
|  |  |  |  | Psychometric quality: -<br>**Quality of evidence:** Low [35] |  |  |  |  |
| Self-Management | Adolescent HIV Self-Management Scale (AdHIVSM) | **study 3:** [47]<br>Methodology quality: Very good<br>Psychometric quality: +<br>**Study 1:**[45]<br>Methodology quality: Inadequate<br>Psychometric quality: -<br>**Quality of evidence:** Very low | **Study 2** [46]:<br>Methodology quality: Very good<br>Psychometric quality: +<br>**Study 3** [47]:<br>Methodology quality: Very good<br>psychometric quality: + | **Study 1:** [45]<br>Methodology quality: Doubtful<br>Psychometric quality: +<br>**Study 2:** [46]<br>Methodology quality: Adequate<br>Psychometric quality: +<br>**Study 3** [47]<br>Methodology quality: Adequate<br>Psychometric quality: +<br>**Quality of evidence:** Low |  |  |  |  |
| Self-compassion | Self-compassion scale (SCS) |  |  | Methodology quality: Doubtful<br>Psychometric quality: +<br>**Quality of evidence:** Low [32] |  |  |  |  |
| Self-Concept | Beck Youth Self-Concept Scale (BYSCS) |  |  | **Study 1** [50]:<br>Methodology quality: Inadequate<br>Psychometric quality: -<br>**Study 2** [51]:<br>Methodology quality: Inadequate<br>Psychometric quality: -<br>**Study 3** [52]:<br>Methodology quality: Moderate<br>Psychometric quality: +<br>**Overall Quality of evidence:** Very low | **Study 1** [50]:<br>Methodology quality: Inadequate<br>Psychometric quality: +<br>**Study 2** [51]:<br>Methodology quality: Inadequate<br>Psychometric quality: ?<br>**Study 3** [52]:<br>Methodology quality: Inadequate<br>Psychometric quality: +<br>**Overall Quality of Evidence:** Very low |  |  |  |
|  | Tennessee Self-concept Scale-2 (TSCS-2) |  |  | **Study 1** [54]:<br>Methodology quality: Doubtful<br>Psychometric quality: +<br>**Study 2** [53]:<br>Methodology quality: Doubtful |  |  | Methodology quality: Doubtful<br>Psychometric quality: -<br>**Quality of evidence:** Very low [54] |  |
|  |  |  |  | Psychometric quality: +<br>**study 3** [55]<br>Methodology quality: Inadequate<br>Psychometric quality: ?<br>**Quality of evidence:**<br>Very low |  |  |  |  |
| Self-efficacy | Self-Efficacy Questionnaire for Children (SEQC) |  |  |  |  |  |  | Methodology quality: Inadequate<br>Psychometric quality: +<br>**Quality of evidence:**<br>Very low [57] |
|  | Self-Efficacy for Managing Chronic Disease 6-Item Scale (SE-6-Xhosa) |  |  | Methodology quality: Inadequate<br>Psychometric quality: +<br>**Quality of evidence:**<br>Very low [58] |  |  |  |  |
|  | Self-efficacy against unwanted sex (SEPOUS) |  |  |  |  |  |  | Methodology quality: Inadequate<br>Psychometric quality: +<br>**Quality of evidence:**<br>Very low [59] |
|  | Self-efficacy for correct condom use (SECCU) |  |  |  |  |  | Methodology quality: Inadequate<br>Psychometric quality: -<br>**Quality of evidence:**<br>Very low [33] |  |
|  | Self-efficacy for negotiating condom use (SENCU) |  |  |  | Methodology quality: Inadequate<br>Psychometric quality: -<br>**Quality of evidence:**<br>Very low[33] |  | Methodology quality: Inadequate<br>Psychometric quality: -<br>**Quality of evidence:**<br>Very low [33] |  |
|  | Self-efficacy to disclose HIV Questionnaire (SEDHQ) |  |  |  |  | Methodology quality: Doubtful<br>Psychometric quality: -<br>**Quality of evidence:**<br>Very low [60] |  |  |
| Self-esteem | Rosenberg<br>Self-esteem<br>Measure<br>(RSEM-10) |  |  | **Study 1** [38]:<br>Methodology<br>quality:<br>Inadequate<br>Psychometric<br>quality: +<br>**Study 2** [64]:<br>Methodology<br>quality:<br>Inadequate<br>Psychometric<br>quality: +<br>**Study 3** [63]:<br>Methodology<br>quality:<br>Inadequate<br>Psychometric<br>quality: +<br>**Study 4** [62]:<br>Methodology<br>quality:<br>Inadequate<br>Psychometric<br>quality: +<br>**Study 5**[42]:<br>Methodology<br>quality: Very<br>good<br>Psychometric<br>quality: +<br>**Study 6**[28]:<br>Methodology<br>quality:<br>Adequate<br>Psychometric<br>quality: +<br>**Study 7** [43] :<br>Methodology<br>quality: Very<br>good<br>Psychometric<br>quality: +<br>**Overall**<br>**Quality of**<br>**evidence:**<br>Moderate | **Study 8**[65]:<br>Methodology<br>quality:<br>Inadequate<br>Psychometric<br>quality: ? | **Study 9** [60]:<br>Methodology<br>quality:<br>Doubtful<br>Psychometric<br>quality: - | **Study 10**[57]:<br>Methodology<br>quality:<br>Doubtful<br>Psychometric<br>quality: -<br>**Quality of**<br>**evidence:**<br>Very low |  |
|  | Modified<br>Rosenberg<br>Self-esteem<br>Measure<br>(RSEM-8) |  |  |  |  |  |  | Methodology<br>quality:<br>Inadequate<br>Psychometric<br>quality: -<br>**Quality of**<br>**evidence:**<br>Very low<br>[59] |
|  | Self-esteem-Hare Area-specific self-esteem scale<br>(HASSES) |  | Methodology<br>quality:<br>Inadequate<br>Psychometric<br>quality: -<br>**Quality of**<br>**evidence:**<br>Very low<br>[39] |  | Methodology<br>quality:<br>Inadequate<br>Psychometric<br>quality: -<br>**Quality of**<br>**evidence:**<br>Very low<br>[39] |  |  |  |
|  | Ad hoc |  |  |  |  |  | Methodology<br>quality: |  |
|  |  |  |  |  |  |  | Inadequate<br>Psychometric<br>quality: +<br>**Quality of**<br>**evidence:**<br>Very low<br>[33] |  |
|  | Ad hoc |  | Methodology<br>quality:<br>Inadequate<br>Psychometric<br>quality: +<br>**Quality of**<br>**evidence:**<br>Very low<br>[38] |  |  |  |  |  |
|  | Ad hoc |  |  |  |  |  |  | Methodology<br>quality:<br>Inadequate<br>Psychometric<br>quality: +<br>**Quality of**<br>**evidence:**<br>Very low<br>[29] |
| Self-worth | Ad hoc |  |  |  |  |  |  | Methodology<br>quality:<br>Inadequate<br>Psychometric<br>quality: +<br>**Quality of**<br>**evidence:**<br>Very low<br>[29] |
| Transcendent | Missoula Vitas<br>Quality of Life<br>Index<br>(MVQOLI)<br>Transcendent<br>Subscale |  |  |  | Methodology<br>quality:<br>Moderate<br>Psychometric<br>quality: +<br>**Quality of**<br>**evidence:**<br>Moderate<br>[66] |  |  |  |

#### Body Appreciation

Body appreciation is defined as “accepting, holding favourable attitudes toward, and respecting the body, while also rejecting media-promoted appearance ideals as the only form of beauty” [27].

##### The Body Appreciation Scale-2 (BAS-2)

The BAS-2 was cited in one study [28]. There was moderate evidence of construct validity. The study was of moderate quality; only the internal consistency of the comparator outcomes was reported.

#### Confidence

Confidence can be defined as the belief in one’s capability to meet the demands of any task [27].

##### Ad-hoc confidence questionnaire

An ad hoc questionnaire measured confidence in a study [29]. There was very low evidence of construct validity; the study was of inadequate quality. Although the outcome was pilot tested and adapted for local use with 10 participants, the outcome measure was not formally validated.

#### Coping

Coping is defined as “strategies, i.e., behaviours, skills, or ways of regulating thoughts and emotions for dealing with stressors [30]. Coping was reported in three studies; all used different outcome measures [31–33].

##### Acceptance of Illness Scale (AIS)

The AIS was cited in one study [32]. There was very low evidence of construct validity. The study was of inadequate quality; only the internal consistency of the comparator outcomes was reported, and the outcome measures were not translated and validated for local use.

##### Ad hoc coping questionnaire

An ad hoc questionnaire was used to measure coping with HIV in one study [33]. There was very low evidence of construct validity. The study was of inadequate quality; no psychometrics of the comparator outcomes were reported, and the outcome measures were not translated and validated for local use.

##### Psychological Adjustment to Illness Scale Self Report (PAIS-SR)

The PAIS-SR was cited in one study [31]. There was very low evidence of construct validity and responsiveness. The study was of inadequate quality; only the internal consistency of the comparator outcomes was reported. Also, the outcome measures were not translated and validated for local use. Lastly, inappropriate tests were used for analysis to measure responsiveness; t-tests were used for a very small sample (N=19).

#### Flourishing

Flourishing can be defined as “a combination of feeling good and functioning effectively and is synonymous with a high level of mental well-being” [34].

##### The Flourishing Well Being Scale (FWBS)

The FWBS was cited in one study [35]. There was moderate evidence of construct validity. The study was of very good quality, and comparator outcome measures were translated and validated in the research setting. However, the study produced null findings; flourishing was equitable for those on Antiretroviral therapy (ART) and those not on ART.

#### Meaningfulness

Meaningfulness can be defined as “the cognizance of order, coherence, and purpose in one’s existence, the pursuit and attainment of worthwhile goals and an accompanying sense of fulfilment” [36].

##### The HIV Meaningfulness Scale (HIVMS)

The HIVMS was cited in one study [32]. There was very low evidence of construct validity. The study was of inadequate quality; only the internal consistency of the comparator outcomes was reported. Further, the outcome measures were not translated and validated for local use.

#### Personal Control

Personal control can be defined as “…a learned repertoire of goal-directed skills that enable humans to act upon their aims, postpone gratification, and overcome difficulties relating to thoughts, emotions, and behaviours” [37].

##### Mastery Scale (MS)

The MS was cited in one study to measure personal control [38]. There was very low evidence of construct validity. The study was of inadequate quality; the outcome measures were not translated and validated for local use. Also, the study sample size was determined based on parameters from a Ghanaian target sample, yet the study compared outcomes across Ghanaian and US participants.

#### Positive Outlook

Positive outlook can be defined as optimism about a great future with or without experiencing adverse events [37].

##### Individual Protective Factors Index (IPFI)

The IPFI was cited in one study [39]. There was very low evidence of internal consistency and construct validity. The study was of inadequate quality. Although the outcomes were translated and adapted for use in Uganda, the investigators used too small a sample for the pilot study (N=15) to establish reliability indices. Further, no additional psychometrics were measured for the adapted outcome measures.

#### Resilience

Resilience is the ability to bounce back from adverse circumstances and maintain optimal mental health functioning [40]. Resilience was evaluated in four studies. One study used the Child Youth Resilience Measure-12 [41], with three studies using the Connor-Davidson Resilience scale (CDRS-10) [35,42,43].

##### Child Youth Resilience Measure-12 (CYRM-12)

The CYRM-12 was cited in one study [41]. There was very low evidence of construct validity. The study was of doubtful quality; the outcome measure was not translated and validated for local use.

##### Connor-Davidson Resilience scale (CDRS-10)

There was high evidence of construct validity of the CDRS-10. All three studies were of very good quality [35,42,43], with the CDRS-10 previously translated and validated in South African adolescents [44].

#### Self-Management

Self-management can be defined as the ability to take necessary steps, including adhering to treatment regimens in managing a condition [45,46].

##### The Adolescent HIV Self-Management Scale (AdHIVSM)

The AdHIVSM was cited in three studies [45–47]. There was high evidence of content validity [47], structural validity [45,47] and internal consistency [46,47]; the studies were of very good quality. There was moderate evidence of construct validity: not all the psychometrics of the comparator outcomes were reported [45,46].

#### Self-compassion

Self-compassion can be defined as “*being open to and moved by one’s own suffering, experiencing feelings of caring and kindness toward oneself, taking an understanding, non-judgmental attitude toward one’s inadequacies and failures, and recognizing that one’s experience is part of the common human experience”* [48].

##### The Self-Compassion Scale (SCS)

The SCS was cited in one study [32]. There was very low evidence of construct validity. The study was of inadequate quality; only the internal consistency of the comparator outcomes was reported, and the outcome measures were not translated and validated for local use.

#### Self-Concept

Self-concept can be defined as how someone perceives and evaluates themselves relative to peers [49]. Six studies evaluated self-concept using the Beck Youth Self-Concept Scale [50–52] and the Tennessee Self-Concept Scale [53–55].

##### The Beck Youth Self-Concept Scale (BYSCS)

The BYSCS was cited in three studies [50–52]. There was very low evidence of construct validity. The three studies were of inadequate quality. Although the secondary outcome measures were translated into the local languages, they were not fully validated. Also, the cross-referenced articles did not contain the validation data as cited by the authors, but rather generic statements on the translation of the outcomes [52,56].

##### Tennessee Self-concept Scale-2 (TSCS-2)

The TSCS-2 was cited in three studies [53–55]. There was very low evidence of construct validity. The three studies were of inadequate quality. Although the secondary outcome measures were translated tools into the local language, they were not validated. Also, the cross-referenced articles did not contain the validation data as cited by the authors [54,55].

#### Self-efficacy

Self-efficacy is defined as self-belief in the capability to execute a specific task regardless of the magnitude of potential obstacles [49]. We analysed six variants of self-efficacy outcome measures reported across five studies [33,57–60].

##### Self-efficacy for correct condom use (SECCU)

An ad hoc questionnaire (SECCU) was used to measure self-efficacy for correct condom use in a single study [33]. There was very low evidence of construct validity. The study was of inadequate quality; no psychometrics of the comparator outcomes were reported, and the outcome measures were not translated and validated for local use.

##### Self-efficacy for negotiating condom use (SENCU)

An ad hoc questionnaire (SENCU) was used to measure self-efficacy for negotiating condom use in a study [33]. There was very low evidence of construct validity. The study was of inadequate quality. No psychometrics of the comparator outcome measures were reported. Also, the outcome measures were not translated and validated for local use.

##### Self-Efficacy Questionnaire for Children (SEQC)

The SEQC was cited in one study [57]. There was very low evidence of construct validity. The study was of inadequate quality. Although the outcomes were translated tools into the local language, they were not formally validated.

##### Self-Efficacy for Managing Chronic Disease 6-Item Scale (SE-6-Xhosa)

The SE-6-Xhosa was cited in one study [58]. There was very low evidence of construct validity. The study was of inadequate quality. There was no transcultural adaptation, and no psychometrics were reported.

##### Self-efficacy to protect oneself from unwanted sex (SEPOUS)

The SEPOUS was cited in one study [59]. There was very low evidence of responsiveness. The study was of inadequate quality; no psychometrics were reported, including the transcultural adaptation of comparator outcome measures.

##### Self-efficacy to disclose HIV Questionnaire (SEDHQ)

The study purpose-built SEDHQ was used in one study [60]. There was very low evidence of responsiveness. The study was of inadequate quality; no scale development and validation process details were provided. Also, only Cronbach’s alpha indices were provided for comparator outcomes.

#### Self-esteem

Self-esteem can be defined as “… as an attitude toward one’s self-based on one’s feelings of worth as a person” [61]. Self-esteem was measured using six different outcome measures, i.e. three ad hoc questionnaires [29,33,38], Hare Area-specific self-esteem scale (HASSES), Rosenburg Self-esteem

Measure (RSEM-10) [28,38,42,43,57,60,62–65] and the Modified Rosenburg Self-esteem Measure (RSEM-8) [39].

##### Ad hoc self-esteem measures

Study purpose-built (ad-hoc) self-esteem questionnaires were used in three studies [29,33,38]. There was very low evidence of construct validity. The studies were of inadequate quality; no details are provided for developing and validating the ad hoc measures.

##### Hare Area-specific self-esteem scale (HASSES)

The HASSES was cited in one study [39]. There was very low evidence of internal consistency and construct validity. The study was of inadequate quality. Although the outcomes were translated and adapted for use in Uganda, the investigators used too small a sample for the pilot study (N=15) to be able to establish reliability indices. Further, no additional psychometrics were measured for the adapted outcome measures

##### Rosenberg Self-esteem Measure (RSEM-10)

The RSEM-10 was used in 10 studies of varying methodological quality. There was moderate evidence for construct validity. The methodological rating ratings were: very good [42,43], adequate [28] doubtful [60,62] and inadequate [38,57,63–65]. The methodological down gradings for the construct validity evaluation studies were mainly due to the lack of reporting of psychometrics; cross-cultural adaptation was not performed in most of the studies [38,57,63,64]. There was high evidence of known-group validity from a single study of very good methodological quality [42].

##### Modified Rosenberg Self-esteem Measure (RSEM-8)

The RSEM-8 was cited in one study [59]. There was very low evidence of responsiveness. The study was of inadequate quality. Two items were omitted from the original RSEM-10, but the transcultural adaptation process details were not provided. Also, no psychometrics were reported, including those of comparator outcome measures.

#### Self-worth

Self-worth can be defined as “…an individual’s evaluation of himself or herself as a valuable, capable human being deserving of respect and consideration” [61].

##### Ad hoc self-worth measure

A study used an ad hoc questionnaire to measure self-worth [29]. There was very low evidence of construct validity. The study was of inadequate quality. Although the outcome was pilot tested and adapted for local use on 10 participants, the outcome measure was not formally validated.

#### Transcendence

Transcendence can be defined as “…a state of existence or perception that is not definable in terms of normal understanding or experience” [66].

##### Missoula Vitas Quality of Life Index (MVQOLI) Transcendence Subscale

The MVQOL was cited in one study [67]. There was moderate evidence of construct validity. The solitary study was of moderate quality; only the internal consistency of the comparator outcomes was reported.

## Discussion

### Overall synthesis

This review sought to identify positive psychological outcomes used in AYALHIV in SSA, map the constructs onto corresponding measures, and critically appraise the identified outcomes’ psychometrics. We gleaned 15 positive psychological constructs, namely: body appreciation, confidence, coping, flourishing, meaningfulness, personal control, positive outlook, resilience, self-management, self-compassion, self-concept, self-efficacy, self-esteem, self-worth and transcendent. Resilience, self-efficacy, and self-esteem were the most measured constructs. Construct validity and internal consistency were the most measured properties, with content and structural validity being the least measured psychometrics. The implications of the individual measurement properties are discussed subsequently.

### Qualitative mapping of positive psychological constructs

Social support was used as an umbrella term to encompass the types of support that occurred at family, peer and community levels. At the interpersonal level, family and peer support were instrumental in assisting adolescents to cope with negative feelings and facilitating belongingness [66,68]. Peer social support assisted ALHIV in achieving the goals of giving and receiving social support, gaining health and relationship advice, adhering to healthcare regimens, learning practical skills and enjoying recreational activities as a group [69]. Social support has also precipitated a positive outlook for life among ALHIV, with adolescents believing that people living with HIV should be allowed to marry and have children if they so desire. Adolescents reporting lack of social support and strained social and interpersonal relations reported neglect, differential treatment, mistreatment [70] and a decreased sense of belonging [17]. The intersection between social support and stigma becomes more obvious at the community level. Adolescents fear disclosing their status because they fear stigmatisation, ridicule, gossip and insult within the school and community [70,71].

Robust self-esteem among adolescents with HIV enabled them to overcome stigma, increase their self-reliance and accept their HIV status [72]. Resilience is shaped by cultural and religious beliefs and the capacity to self-reflect and face adverse conditions [72,73]. Resilient adolescents had greater life satisfaction [73], and were acceptive of their live circumstances. Resilience was vital for young persons to cope with their realities [72] and muster the courage to face possible stigma [70]. Among adolescents, disclosure to others was not always based on choice; the process is emotional and complex, with uncertain outcomes [74]. However, disclosure positively influenced adherence and retention to care health improvement and enabled social participation [75].

### Structural validity

Despite the wide use of positive psychological measures, the evidence for structural validity was limited. Except for the Adolescent HIV Self-Management Scale [32], all outcome measures analysed were developed in high-income settings but were not properly translated and validated before use in different contexts. Robust transcultural adaptations are essential for preserving structural validity, an essential psychometric property [76–78]. Structural/factorial validity measures the extent to which items measure the latent constructs purportedly measured by a specific outcome measure [76]. For example, the Connor-Davidson Resilience scale was the most commonly used resilience outcome measure, applied in three of the five studies of resilience [35,42,43], yet none of these studies evaluated structural validity. Also, the Rosenburg Self-esteem Scale, was used in ten studies evaluating self-esteem [38,42,43,57,60,62–64] but no study evaluated its structural validity.

Further, an outcome measure may perform differently when applied to two different geographical regions in the same country owing to sociocultural and linguistic differences. For instance, the Flourishing Scale exhibited measurement invariance/diffrential item functioning in South African university students [79]. Three of the eight items performed differently across the study’s four languages, i.e. English, Afrikaans, Sesotho and Setswana [79]. Robust transcultural translations and adaptions, including structural validity assessment, are essential before using an outcome measure with seemingly high psychometric robustness in another country. Psychometric performance in another country can never be assumed.

During the analysis, we observed a trend of snowball citations, i.e. the tendency to cite previously published studies to justify the validity of applied outcome measures [80]. Snowball referencing is problematic as the actual measurement properties of most positive psychological outcomes remain elusive. For example, a Kenyan study explored the construct validity of the Rosenberg Self-esteem Scale by investigating the correlates of self-esteem to self-efficacy in HIV treatment adherence as measured by the ART Adherence Self-efficacy (HIV-ASES) [64]. The study by Gitahi-Kamau et al. (2022) cites a previous validation study performed in the US as evidence of the psychometric robustness of the HIV-ASES [81]. However, the HIV-ASES did not undergo transcultural adaptation and validation before use in Kenya; this may lead to measurement bias.

### Construct validity

Construct validity is the extent to which scores on two outcomes correlate [76]. Sufficient evidence of structural validity is a pre-requisite for construct validity [76]. In this review, construct validity was the most measured psychometric, with most tools showing evidence of moderate to high collective robustness. The high construct validity evidence across outcome measures may indeed imply that the outcomes were measuring what they were supposed to measure. However, the lack of structural validity may “invalidate” evidence of construct validity robustness [76]. This contradiction (lack of structural validity) poses a measurement error dilemma, as most outcomes still performed satisfactorily. Most of the outcomes had positive ratings regarding the quality of construct validity. The Pearson correlation coefficient was the most applied bivariate correlation index. The robustness of the Pearson correlation is a function of normality and sample size. None of the studies reported normality indices. However, most studies recruited samples ≥100; with a larger sample size, weak correlations are likely to show statistical significance. That said, the correlations were in the moderate ranges, with only two studies yielding very strong correlations, i.e., R≥0.8; this may downgrade the overall evidence of construct validity. Nevertheless, building on the strong construct validity evidence across outcomes, there is a need for solid efforts for proper validation i.e. measuring both structural and construct validity to ensure measurement equivalency to facilitate cross-cultural comparisons [76–78].

### Responsiveness

Responsiveness is the ability of an outcome measure to detect change over time [76]. There was insufficient evidence of responsiveness across the outcomes. Despite pilot testing and adaptation, none of the studies evaluating responsiveness formally validated outcomes before use [29,57,59]. In some instances, no psychometrics were provided [59]. For example, a randomised controlled trial evaluated the effectiveness of a peer-led intervention in improving linkage to and retention in care, adherence to ART and psychosocial well-being among adolescents living with HIV in rural Zimbabwe [29]. The study applied an ad-hoc questionnaire measuring confidence, self-esteem, and self-worth as secondary outcomes. Although the positive psychological outcome measures were translated into the native language and pilot tested, the tools were not formally validated. The study demonstrated intervention effectiveness on the primary outcome (viral load suppression) and positive psychological constructs. It is reasonable to scale up the intervention at the clinical level. However, this may be problematic when inferring the intervention effects on the measured positive psychological outcomes. Using unvalidated outcomes may distort intervention effect sizes, which may lead to incorrect conclusions. More efforts are required to ensure that positive outcomes are adequately validated, with norms or cut-off scores identified before the measures are used for intervention(s) evaluation.

### Reliability

Reliability measures the extent of stability and reproducibility of outcomes assuming constancy in extraneous variables [76,82]. The Cronbach alpha was the most cited reliability index, with overall evidence of reliability in the moderate to high range. As observed in previous reviews, the Cronbach Alpha was inappropriately used in most studies as an indicator of psychometric robustness [80,83,84]. There were instances where outcome measures were adapted and translated, with the Cronbach alpha cited as evidence of reliability and validity. The Cronbach alpha measures the degree of connectedness of items; it is neither a true indicator of internal consistency, a form of reliability, nor validity [76,82,85]. Evaluation of Cronbach alpha is not a substitute for full validation. Compared to other forms of reliability, such as test-retest reliability, and split-half reliability indexes, the Cronbach alpha is the “least desired/robust” reliability indicator [82,85]. Although most outcomes yielded high Cronbach alphas, there is a need for properly designed and fully-powered psychometric evaluation studies. For instance, structural validity must be established before evaluating internal consistency and construct validity [76].

### Clinical and research utility

All but three outcome measures were available free of charge; this increases the utility of the identified positive psychological outcome measures. Most of the outcomes were rated on 4- or 5-point Likert scales, with some using 7-point Likert scales. Consideration needs to be made during transcultural adaptations to ensure age- and developmental-appropriate adaptations. For example, it essential to decrease the number of response options to increase the feasibility of use in AYALHIV. For instance, HIV-related neurological impairment can decrease AYALHIV cognitive capabilities. In Africa, more common lower levels of education, and HIV itself, can impede school attendance and learning in this context. A previous validation study in Uganda had to collapse seven response options to five in addition to using visual cues as participants had difficulties understanding the original scoring instructions [86]. Cultural and linguistic differences must be accounted for to ensure equivalency between the original and source languages [77]. Robust transcultural translations and adaptations are critical, given that most of the outcomes gleaned were from high-income countries. Most of the outcome measures were brief; this decreases respondent burden and increases the feasibility of research and routine use for evidence-based care. Also, most of the outcomes were generic; this allows comparisons across conditions and settings and could be applicable for use in other chronic conditions. However, very few tools had established cut-off points; this makes comparisons across studies and contexts difficult. Overall, most of the tools had a high utility for routine use, given that most were generic, brief, had fewer response options and were available at low or no cost [25].

### Limitations

A significant limitation of the current review is that most of the studies analysed were not primarily psychometric evaluation studies. As such, the odds of high risk of bias (RoB) ratings were great given that the COSMIN checklist, which we utilised to evaluate methodological quality, was primarily designed to appraise psychometrics evaluation studies. For example, there was poor evidence of responsiveness across the outcome measures analysed. None of the analysed studies were primarily designed to evaluate responsiveness, rather, we analysed results from interventional studies to evaluate responsiveness. The COSMIN checklist is considered a “gold standard” for RoB evaluations but has limitations; it overtly gravitates to the stringent spectrum of psychometric RoB checklists [87]. However, we utilised multiple methods to ensure fair judgements per study. For instance, we contacted authors to get information essential for RoB ratings, which may not have been published to avoid reporting bias. Also, we had consensus meetings to synthesise all findings, as the first round of RoB ratings had yielded poor ratings for most outcomes. Due to resource limitations, we only analysed peer-reviewed articles published in English; this may have introduced language bias. Nevertheless, applying the PRISMA guidelines throughout increases the robustness of the review findings despite the inevitable methodological pitfalls.

## Conclusion

We identified 15 positive psychological constructs applied in AYALHIV in SSA: body appreciation, confidence, coping, flourishing, meaningfulness, personal control, positive outlook, resilience, self-management, self-compassion, self-concept, self-efficacy, self-esteem, self-worth and transcendent. Of the identified outcome measures, the RSEM-10, Missoula Vitas Quality of Life Index Transcendent Subscale, the Adolescent HIV Self-Management Scale, Connor-Davidson Resilience scale, Flourishing Well-Being Scale, and the Body Appreciation Scale-2 had moderate to high evidence of psychometric robustness. Few studies performed complete validations; thus, evidence for psychometric robustness was fragmented. However, this review demonstrates the initial evidence of the feasibility of positive psychological outcomes for use in AYALHIV in low-resource settings. Instead of creating new outcomes, authors are advised to leverage the existing outcomes, adapt them for use, and, if appropriate, strive to maintain the factorial structure to facilitate comparisons. Lastly, validating composite positive psychological outcomes should be considered for transcultural adaptations, given the variable psychometric performance across constructs and measurement properties.

## Data Availability

We will be extracting data from published articles no datasets will be generated.

## Abbreviations

AdHIVSM: Adolescent HIV Self-Management Scale
AIDS: Acquired Immunodeficiency Syndrome
AIS: Acceptance of Illness Scale
ALHIV: Adolescents Living With HIV
ART: Anti Retroviral Therapy
AYALHIV: Adolescents and Young Adults Living With HIV
BAS-2: Body Appreciation Scale
BYSCS: Beck Youth Self Concept Scale
CDRS: Connor Davidson Resilience Scale
CINAHL: Cumulative Index of Nursing and Allied Health Literature
COSMIN: COnsensus-based Standards for the selection of health Measurement Instruments
CYRM-12: Child Youth Resilience Measure
FWBS: Flourishing Wellbeing Scale
GRADE: Grading of Recommendations Assessment, Development, and Evaluation
HASSES: Hare Area Specific Self Esteem Scale
HIV: Human Immunodeficiency Virus
HIV-ASES: HIV ART Adherence Self Efficacy Scale
HIVMS: HIV Meaningfulness Scale
HRQoL: Health-Related Quality of Life
IPFI: Individual Protective Factors Index
MeSH: Medical Subject Headings
MS: Mastery Scale
MVQOLI: Missoula Vitas Quality of Life Index Transcendence Subscale
PAIS-SR: Psychological Adjustment to Illness
Scale: Self Report
PMHI: Positive Mental Health Interventions
PRISMA: Preferred Reporting Items of Systematic Reviews and Meta-Analyses
PRISMA-P: Preferred Reporting Items of Systematic Reviews and Meta-Analyses Protocol
PRISMA-ScR: Preferred Reporting Items of Systematic Reviews and Meta-Analyses extension for Scoping Reviews
PROSPERO: Prospectively Registered Systematic Reviews with a health related Outcome
RoB: Risk of Bias
RSEM: Rosenberg Self Esteem Measure
SCS: Self Compassion Scale
SECCU: Self Efficacy for Correct Condom Use
SEDHQ: Self Efficacy to Disclose HIV Questionnaire
SENCU: Self Efficacy for Negotiating Condom Use
SEPOUS: Self Efficacy to Protect Oneself from Unwanted Sex
SEQC: Self Efficacy Questionnaire for Children
SE-6-Xhosa: Self Efficacy for managing chronic disease 6 item scale Xhosa Version
SSA: Sub Saharan Africa
TSCS-2: Tennessee Self-Concept Scale
TWCF: Templeton World Charity Foundation
YEP: Youth Expert Panel

## Declarations

### Ethics approval and consent to participate

Not applicable.

### Consent for publication

Not applicable.

### Availability of data and materials

Not applicable as we reviewed published studies. However, all relevant data/materials, including data collection tools, were submitted as supplementary files.

## Supporting information

S1 Table : Preferred Reporting Items for Systematic reviews and Meta-Analyses extension for Scoping Reviews (PRISMA-ScR) Checklist

S2 Table: PRISMA Checklist

S3 Table : CINAL search strategy

S4 Table : Operational definitions of psychometric properties

S5 Table: Updated criteria for good measurement properties

S6 Table : GRADE checklist-best evidence synthesis

S7 Table : Study selection

S8 Table : Qualitative mapping of psychological constructs S9 Table : Outcomes utility

